# Immune Checkpoint Response Profiles and Resistance Mechanisms in NSCLC Revealed by Circulating Extracellular Vesicle Proteomics

**DOI:** 10.64898/2026.05.25.26354042

**Authors:** C. Taylor, M. Davey, E. P. Allain, A. Sabir Cheema, N. Crapoulet, N. Finn, M. Abdelsalam, R. J. Ouellette

## Abstract

**Background:** Immune-oncology has revolutionized cancer treatment, but some patients fail to benefit due to primary resistance and tumour-immune evasion. Extracellular vesicles (EVs) are secreted by both tumour and immune cells and mediate communication between cancer cells and the immune system. Our study used proteomic profiling of circulating EVs collected from NSCLC patients treated with immune checkpoint inhibitors (ICI) to identify predictive biomarkers of response as well as immune evasion mechanisms related to treatment resistance.

**Methods:** EVs were isolated from plasma collected prior to ICI treatment using peptide-affinity purification and high-throughput proteomics was performed using Proximal Extension Assay. Differentially expressed EV proteins between durable (DR) and non-durable responders (NDR) were identified and evaluated using Cox proportional hazards regression, survival analysis, sex-stratified analysis, as well as pathway and network analysis.

**Results:** Proteomics analysis identified 116 differentially expressed EV proteins between DR and NDR. NDR was characterized by enrichment of inflammatory, angiogenic, and immune-suppressive EV proteins, such as IL1RL1, TFRC, IL6ST, galectins, TNF superfamily death receptors, chemokines, and PCSK9. Pathway analysis revealed enrichment of angiogenesis, chemotaxis, ECM remodeling, and neutrophil degranulation associated with poor progression-free survival (PFS). In contrast, DR to ICI treatment was associated with EV proteins related to T- and B-cell activation and adaptive immunity. Sex-related differences in abundance and association with PFS was observed for certain EV proteins, including IL1RL1 and TFRC. A six protein EV model (IL1RL1, TFRC, ERI1, CCN5, IGFBPL1, and TNFRSF13C) demonstrated good prognostic performance for identifying NDR (AUC = 0.907) and stratified patients into three discrete risk groups.

**Conclusions:** High-plex EV proteomics revealed biologically coherent tumour-immune signaling programs that are associated with ICI treatment resistance. Profiling circulating EVs may improve our understanding of EV-mediated immune evasion mechanisms and identify protein signatures that reflect the tumour immune microenvironment and predict response to immune checkpoint blockade.

- **What is already known on this topic** – Immune checkpoint inhibitors (ICIs) have improved outcomes in NSCLC, but many patients exhibit primary resistance due to tumour–immune evasion. Extracellular vesicles (EVs) mediate tumour–immune communication and are a promising source of circulating biomarkers, though their proteomic role in predicting ICI response remains poorly defined
- **What this study adds** – Our findings suggest that EVs provide a minimally invasive, systems-level readout of tumour–immune dynamics and can simultaneously serve as a source of predictive biomarkers and mechanistic insight into treatment resistance.
- **How this study might affect research, practice or policy** – This work may inform biomarker-driven patient stratification, guide future therapeutic strategies, and encourage incorporation of EV proteomics and sex-specific analyses in clinical research.

## INTRODUCTION

Precision medicine has created a rapidly changing landscape in the treatment of non-small cell lung cancer (NSCLC) with the introduction of targeted therapies against oncogenic drivers and more recently, by the widespread adoption of immune checkpoint blockade (ICB). Immune checkpoint inhibitors (ICIs) disrupt immune checkpoint proteins enabling tumour cells to avoid T cell-mediated immune surveillance, eliciting durable responses for some patients. Inhibitors against programmed death protein 1 (PD-1), it’s ligand (PD-L1), or cytotoxic T lymphocyte antigen 4 (CTLA-4), administered as monotherapy or in combination with chemotherapy, now represent the standard of care for NSCLC patient’s ineligible for targeted therapies.

Despite major advances in immune checkpoint blockade, relatively little is known about the biological determinants of individual patterns of treatment response or failure, particularly at the level of host–tumor interactions that pre-configure benefit or resistance. Sex is one of the clearest but still underexplored variables in this context. Meta analysis of patients treated with ICI have revealed men derive greater benefit form anti-PD-1/PD-L1 monotherapy, particularly in PD-L1+ tumours, while women show greater survival benefit when treated with PD-L1 inhibitors with combination chemotherapy^1^. Complex interactions among sex-linked genes, behaviour, and particularly sex hormones, likely drive these sex-related differences in response to ICB.

Tumour PD-L1 expression is currently used to stratify patients for ICI therapy, however it is not a reliable predictor of ICI response or survival benefit^2^. Although the introduction of ICIs has greatly improved overall survival in NSCLC patients, many patients fail to achieve a durable response due to either primary or acquired immunotherapy resistance^3^. Response to ICI therapy requires the recognition and presentation of cancer antigens by antigen presenting cells as well as a robust T-cell mediated response and immune cell-mediated tumour killing with the tumour microenvironment (TME). An immunosuppressive TME, characterized by increased presence of M2-polarized macrophages, myeloid-derived suppressor cells (MDSCs), and neutrophils, together with exclusion of B cells and cytotoxic effector cells, such as CD8+ T cells and natural killer (NK) cells, can further contribute to primary ICI resistance.

EVs constitute a heterogenous population of membrane-bound vesicles and that are secreted from all cell types, both from the endosomal system and shed from the plasma membrane and therefore frequently display a functionally rich repertoire of membrane-associated proteins and signaling receptors that are reflective of both the type and status of their cell of origin^4^. EVs released by malignant cells can exert pro-tumoral effects by supporting tumour survival and immune evasion through suppression of T cell and NK cell function, promoting myeloid, neutrophil, and regulatory T cell (Treg) recruitment, and inducing the differentiation of immune-suppressive tumour-associated macrophages (TAMs) and neutrophils^5^. EVs secreted by immune cells enables bi-directional crosstalk between the tumour and immune system. Immune-derived EVs exert both short and long-range immune modulating effects, impacting diverse pathways associated with inflammation, immune-suppression or activation, and TME remodeling, in a context-dependent manner reflective of the signaling molecules received by their cell(s) of origin^4^. Liquid biopsies have become increasingly important due to their ability to identify tumour-derived biomarkers, cell-free DNA, and EVs that support patient monitoring during treatment to identify recurrences and therapeutic escape. Circulating EVs are enriched in immune-oncology relevant proteins including immune checkpoint proteins and immune-modulating proteins such as cytokines, chemokines and growth factors, making them a promising avenue for biomarker discovery in personalized cancer immunotherapy^6^. EVs bearing PD-L1 are increasingly recognized as an important mechanism of both local and systemic immune suppression and immunotherapy resistance and have shown promise as predictive marker for ICI response^5^.

Predictive biomarkers of ICI response are critical to efforts to optimize immunotherapy treatments. A robust biomarker response panel could improve response rates by identifying patients most likely to benefit from treatment, while simultaneously reducing unnecessary toxicity and healthcare costs by directing patients unlikely to respond towards alternative treatment options.

In this study, we used high-throughput proteomics to profile the plasma EV proteome to improve our understanding of molecular drivers of response to ICB. Given the inherent sex-related differences in immune modulation, we also examined sex-specific variation within the EV proteome. This exploratory pilot study included patients with metastatic NSCLC who initiated treatment with ICI therapy between 2018 and 2024. Baseline plasma EVs were profiled using high-throughput proteomics to identify EV protein features associated with either durable response (DR) or non-durable response (NDR) to ICB. Cox proportional hazards regression and survival analysis was used to identify plasma EV proteins correlated to progression-free survival (PFS). Using this approach, we established a novel circulating EV proteomic signature with good predictive performance for identifying patients unlikely to derive durable clinical benefit from ICI therapy.

## METHODS

### Patient Cohort

EDTA plasma samples from a cohort of 114 patients with metastatic non-small cell lung cancer (NSCLC) who began treatment with immune checkpoint inhibitors between 2017 and 2025 were obtained from the CHU Dumont Biobank (Moncton, New Brunswick, Canada). Whole blood was processed into plasma by centrifugation at 1,500xg for 15 minutes within 30 minutes of collection, to minimize ex vivo platelet activation which can alter EV composition and number, and stored at -80ºC. The study was conducted in accordance with the Declaration of Helsinki with approval from the research ethics boards of the Vitalité and Horizon Health Networks. Patients whose disease did not progress, as determined by imaging, for at least 12 months after initiation of treatment with anti-PD-1/anti-PD-L1 were classified as durable responders (DR) whereas patients who experienced disease progression in <12 months were deemed to be non-durable responders (NDR). All patients received at least two cycles of anti-PD-1/PD-L1 therapy and were followed for a minimum of 1 year. Progression-free survival was defined as the duration, in days, between the date of the first ICI treatment dose and the date of either disease progression or death due to any cause. Patients who terminated treatment in <1 year due to intolerance or who were lost to follow-up were deemed to be non-evaluable.

### EV Isolation and Lysis

EVs were isolated from freshly thawed plasma using peptide affinity precipitation with Vn96 peptide^7^. Plasma was pre-cleared by centrifugation at 3,000xg for 15 minutes at 4ºC for 15 minutes, transferred to a new tube and then diluted 1:1 in PBS and 0.25% Protease Inhibitor Cocktail III (ThermoFisher Scientific, Mississauga, ON). Vn96 peptide was solubilized at 2.5 mg/mL in 0.625X PBS + 0.05% Pro-Clin300, added at a final concentration of 0.05 mg/mL in 1 mL of diluted plasma, and incubated for 1 hour with end-over-end rotation. The samples were centrifuged at 17,000xg for 15 minutes at 4ºC to precipitate the EV/peptide complexes and the pellets were washed three times with PBS. The EV pellets were lysed in a modified RIPA buffer [50 mM Tris-HCl (pH 7.4), 150 mM NaCl, 1 mM EDTA, 1 % Triton X-100, 0.1% sodium deoxycholate, and 1x protease inhibitor cocktail)] followed by incubation for 10 minutes with shaking at 1,000 rpm and storage at -20ºC. After thawing the EV lysates, insoluble material was removed by recovering the supernatant following centrifugation at 10,000xg for 5 minutes. The protein concentration was measured (Pierce BCA Protein Assay Kit; ThermoFisher Scientific) and the samples were diluted to a final concentration of 0.5 mg protein/mL using the modified RIPA buffer.

### Proximity Extension Assay

Plasma EV proteomic analysis was performed by Olink® Proximity Extension Assay (PEA) (Olink Proteomics AB, Uppsala, Sweden) using the Explore 3072 assay with coverage of 2941 individual proteins. In brief, the assay was performed by incubating pairs of oligonucleotide-linked antibodies with 20 µL of EV protein lysate at a concentration of 0.5 mg/mL (10 µg total protein) and the assay was performed according to Olink’s protocol for EVs, sequenced by Next Generation Sequencing and the counted reads were translated into normalized protein expression (NPX). Data were normalized using Olink’s standard workflow, including internal control adjustment, plate control normalization, and intensity-based normalization across randomized plates. Samples or assays which failed QC were removed from the analysis.

### Statistics and Bioinformatics

As the purpose of this study was exploratory and hypothesis-generating rather than confirmatory and given the large number of analytes profiled using the Explore 3072 platform, statistical significance was evaluated using unadjusted p-values. False discovery rate (FDR) correction was calculated using the Benjamini–Hochberg procedure and used for reference but was not used to define statistical significance. Lists of differentially expressed proteins, Principal Component Analysis (PCA) plots, and heatmaps were generated using Qlucore Omics Explorer software (Qlucore, Sweden) and comparisons between two groups were performed using the Mann–Whitney U test. Time-to-event analyses were performed using Kaplan–Meier survival curves, with between-group differences assessed using the log-rank test. Cox proportional hazards regression was used to estimate hazard ratios (HR) and the association of profiled proteins with PFS. Pathway enrichment analysis was performed using the Metascape web tool and used to identify pathways with significant fold changes (FDR<0.05). Multiple logistic regression was performed to estimate odds ratios and β-coefficients using online statistical software (Stats.Blue). A stepwise variable removal approach was applied to derive the final model. Predicted probabilities were calculated using the logistic function: logit(p)=β0+β1X1+β2X2+⋯+βkXk. Receiver operating characteristic (ROC) analysis was used to evaluate the discriminatory performance of selected models, and the area under the curve (AUC) was calculated as a summary measure of classification performance. R version 4.5.2 (R Foundation for Statistical Computing, Vienna, Austria) was used to perform optimal cut point generation, K-M analyses, Cox regression, ROC analyses, and AUC calculations using ggplot2, survival, survminer, and pROC packages. Protein-protein interaction (PPI) networks were explored using the STRING version 12.0 database with a 0.7 confidence interaction score^8^.

## RESULTS

### Proteomic profiling reveals differences in levels of circulating EV proteins between DR and NDR

With the goal of exploring the proteome of plasma EVs to identify protein signatures that are associated with resistance to ICI, we obtained plasma collected from patients with advanced NSCLC prior to the initiation of treatment. The baseline characteristics of the patient cohort (Table. 1) reflect a population that had not previously been treated with ICI and predominantly received first-line therapy with anti-PD1 therapies including pembrolizumab (n=90, 84.9%) and nivolumab (n=12, 11.3%). A total of 106 patients were evaluable for treatment response and included 57 (53.8%) male and 49 (46.2%) female patients. Disease progression that occurred within the first 12 months of treatment was classified as non-durable response (NDR) and made up 60.4% (n=64) of evaluable patients. Median PFS was 152.5 days for NDR and 698 days for DR (HR_NDR_ = 11.1, 95% CI 6.8-18.0; p<0.0001), however sex was not a significant risk factor for NDR.

**Table 1:** Clinicopathological characteristics of patients with non–small cell lung cancer (NSCLC) treated with anti–PD-1 or anti–PD-L1 therapy, stratified by non-durable responders and responders. Baseline demographic, clinical, and molecular variables are shown, with statistical comparisons between groups. Continuous variables are presented as median ± S.D. and categorical variables as number (percentage). P values indicate differences between non-durable responders and responders. Abbreviations: S.D. = Standard differentiation; ICI = Immune checkpoint inhibition; NDR = Non-Durable Response; DR = Durable Response.

| **Patients and clinical characteristics** | **No Durable Response (n=63)** | **Durable Response (n=43)** | **p value** |
| --- | --- | --- | --- |
| **Median age at start of treatment with ICI (years)** | **71 ± 8.0** | **67 ± 9.3** | **0.002** |
| Males, n (%) | 38 (60.3%) | 19 (44.2%) | 0.105 |
| Females, n (%) | 25 (39.7%) | 24 (55.8%) |  |
| **TMN staging** |  |  |  |
| I/II | 1 (1.6%) | 2 (4.7%) | 0.400 |
| III/IV | 60 (95.2%) | 39 (90.7%) |  |
| Missing data | 2 (3.2%) | 2 (4.7%) |  |
| **First line treatment** |  |  |  |
| Yes | 45 (71.4%) | 33 (76.7%) | 0.760 |
| No | 8 (12.7%) | 7 (16.3%) |  |
| Missing data | 10 (15.9%) | 3 (7.0%) |  |
| **ICI treatment** |  |  |  |
| Pembrolizumab (anti-PD1) | 53 (84.1%) | 37 (86.0%) | 0.685 |
| Nivolumab (anti-PD1) | 8 (12.7%) | 2 (4.7%) |  |
| Nivolumab (anti-PD1) + Ipilimumab (anti-CTLA4) | 0 (0%) | 2 (4.7%) |  |
| Durvalumab (anti-PDL1) | 2 (3.2%) | 2 (4.7%) |  |
| **PDL1 TPS** |  |  |  |
| PDL1 TPS <1% | 16 (25.4%) | 9 (20.9%) | 0.365 |
| PDL1 TPS ≥1% but <50% | 13 (20.6%) | 17 (39.5%) |  |
| PDL1 TPS ≥ 50% | 26 (41.3%) | 10 (23.3%) |  |
| Missing data | 8 (12.7%) | 7 (16.3%) |  |
| **kRAS mutations in tumor tissue** |  |  |  |
| kRAS mutation | 17 (27.0%) | 14 (32.6%) | 0.920 |
| No kRAS mutation | 28 (44.4%) | 22 (51.2%) |  |
| Missing data | 18 (28.6%) | 7 (16.3%) |  |
| **Median progression-free survival (days)** | **152.5** | **698** | **<0.0001** |
| **Median overall survival (days)** |  |  |  |

The Olink Explore 3072 PEA platform was used for proteomic profiling of plasma EV protein lysate isolated from lung cancer patients prior to initiation of treatment with ICIs. A total of 2404 proteins (84%) were detected in >50% of samples. PCA plots were used to assess sample and assay quality using the normalized matrix. Features that clustered distinctly away from the main cluster of assays on the first two principal components were considered outliers and removed from downstream analysis (Supp. Fig. S1). 784 (27.3%) of the assays were excluded on this basis leaving a total of 2090 assays and 106 samples remaining in the final analysis.

To determine whether interrogating the plasma EV proteome can yield predictive biomarkers of ICI response, differences in levels of circulating EV proteins between DR and NDR to ICI at baseline were evaluated (Fig. 1). After excluding non-significant proteins (p>0.01), PCA revealed partial separation of NDR and DR patient clusters (Fig. 1A) and 116 differentially expressed (DE) circulating EV proteins were identified (Fig. 1B-D). Eighty proteins were upregulated in NDR compared to DR, notably proteins related to inflammation and immune signaling including IIL1RL1 (ST2), IL6ST (gp130), CD177, CHI3L1, and growth differentiation factor 15 (GDF15) (Fig. 1C,D). Additionally, iron (TFRC) and lipid metabolism (PCSK9) proteins were also found to be elevated in NDR. Thirty-six proteins were found to be elevated in DR, including CD5, CD27, CD22, SIT1, as well as the T cell co-stimulatory protein CD80. Collectively, this differential expression pattern points to a Th2-type immune signature linked to NDR whereas DR is characterized by a T cell and B cell-associated immune signature.

**Figure 1:**
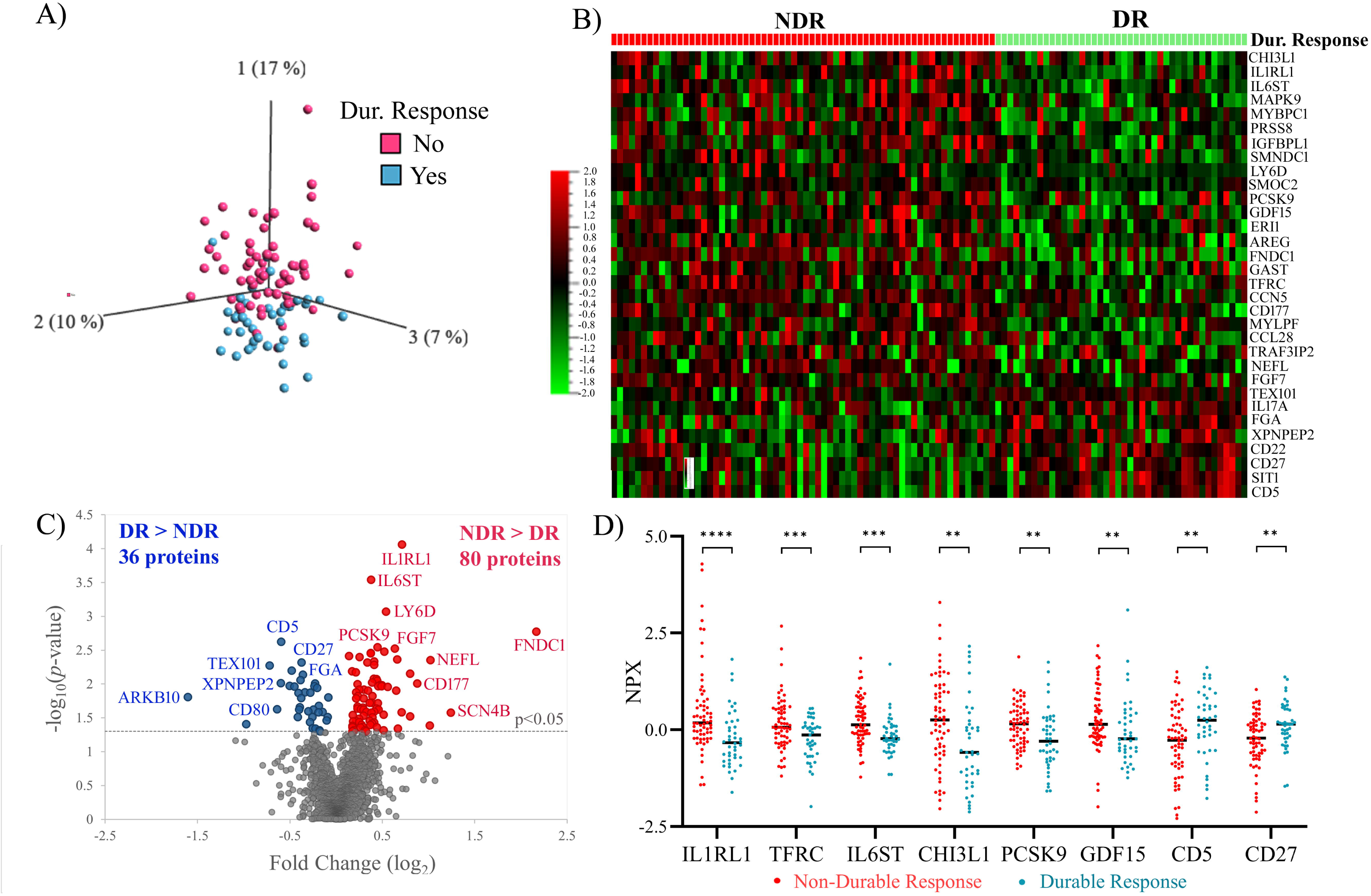
Circulating EV proteomics analysis investigating differences between durable and non-durable response/non-responders to immune checkpoint inhibition therapy in NSCLC patients. (A) Principal component analysis comparing patients who experienced non-durable response to immune checkpoint inhibition to patients who experienced a durable response after applying statistics (p≤0.01). (B) Heatmap depicting top 25 differentially expressed proteins between non-durable responders (NDR) and durable responders (DR) to ICI therapy. (C) Volcano plot comparing circulating EV proteins that are increased in patients who experienced non-durable response to proteins increased in patients who experienced durable response. Boxplot showing differences in mean expression between NDR and DR for selected proteins. Dur. = Durable; NDR = Non-Durable Response; DR = Durable Response; NPX = normalized protein expression. * p<0.05, ** p<0.01, ***p<0.001, ****p<0.0001.

### Sex-disparities were observed in EV proteome at baseline

The DE of circulating EV proteins at baseline between males and females in the cohort was also explored (Fig. 2). Analyzing differences in protein levels between DR and NDR in males and females separately appeared to tighten the clustering of NDR and DR in the PCA (Fig. 2A,B), suggesting the influence of sex-dependent biology on response-associated protein signatures. As observed in the total patient cohort, many of the up-regulated proteins in males were related to immune signaling (e.g. IL1RL1, CD276, PCSK9) and tumour progression (FNDC1, GAST) (Fig. 2C,D,F). FNDC1^9^, which was one of the most highly differentially expressed proteins in the full NDR cohort, was found to be elevated only in male NDR patients and not females (Supp. Fig. S2). Likewise, both IL1RL1 and the immune checkpoint protein CD276 were found to be significantly elevated only in male NDR patients (Fig. 2F). Female NDR patients were also enriched in tumour-related inflammatory markers, including LY6D, GDF15, TFRC, CEACAM19, FGF7, and FGF16 (Fig. 2C,E,F). In contrast, the anti-angiogenic growth factor VEGB was significantly associated with DR in males but not females (Fig. 2C,F). Conversely, TFRC and CD5, which were identified in the full cohort, were found to be much more strongly associated with NDR or DR, respectively, in females. Likewise, CEACAM19 and KIR3DL2, were elevated only in female NDR patients. Therefore, disparities in pre-treatment plasma EV protein signatures correlated with response in a sex-dependent manner, suggesting that sex-related differences in pathways underlying response to ICB may exist.

**Figure 2:**
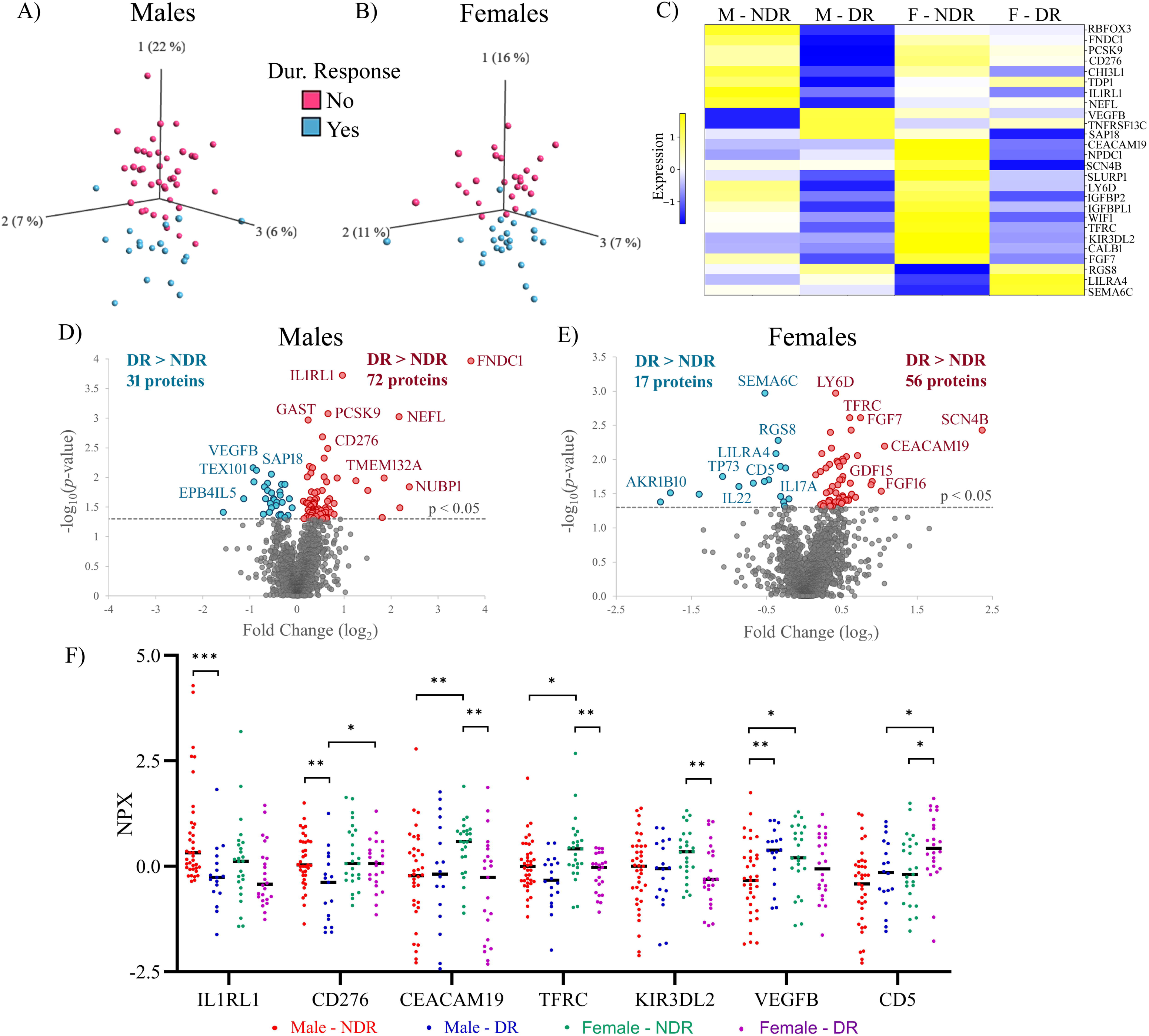
Circulating EV proteomics analysis investigating sex-related differences between durable and non-durable/non-responders to immune checkpoint inhibition of NSCLC patients. Principal component analysis comparing (A) male or (B) female patients who experienced non-durable or non-response to immune checkpoint inhibition to patients who experienced a durable response after applying statistics (p≤0.02). (C) Heatmap depicting mean NPX between top differentially expressed proteins between male and female non-durable and durable responders to ICI therapy. Volcano plot comparing circulating EV proteins that are increased in (D) male or (E) female patients who experienced non-durable response to proteins increased in patients who experienced durable response. (F) Boxplot showing sex-related differences in mean expression between NDR and DR. Dur. = Durable; M = Males; F = Females; NDR = Non-Durable Response; DR = Durable Response; NPX = normalized protein expression. * p<0.05, ** p<0.01, ***p<0.001).

### EV-associated TNF superfamily death receptors and galectins and enrichment of immune suppressive pathways are associated with higher risk of progression

EV proteins associated with PFS were identified using Cox proportional regression analysis (Fig 4A). Approximately 200 EV proteins were found to be significantly associated with risk of shorter PFS, including inflammatory proteins, death receptors and immune checkpoint proteins involved in tumour-immune interactions. TNF superfamily members included TNFRSF6B (DcR3), the TRAIL receptors TNFRSF10A/B and the TWEAK receptor, TNFRSF12A (Fig 4A; Supp. Table S1). The lectins, galectin-1 (Fig. 3A), galectin-3 and galectin-4 which are known to be enriched on tumour-derived EVs^10^, were also correlated to poor response (Supp. TableS1).

**Figure 3:**
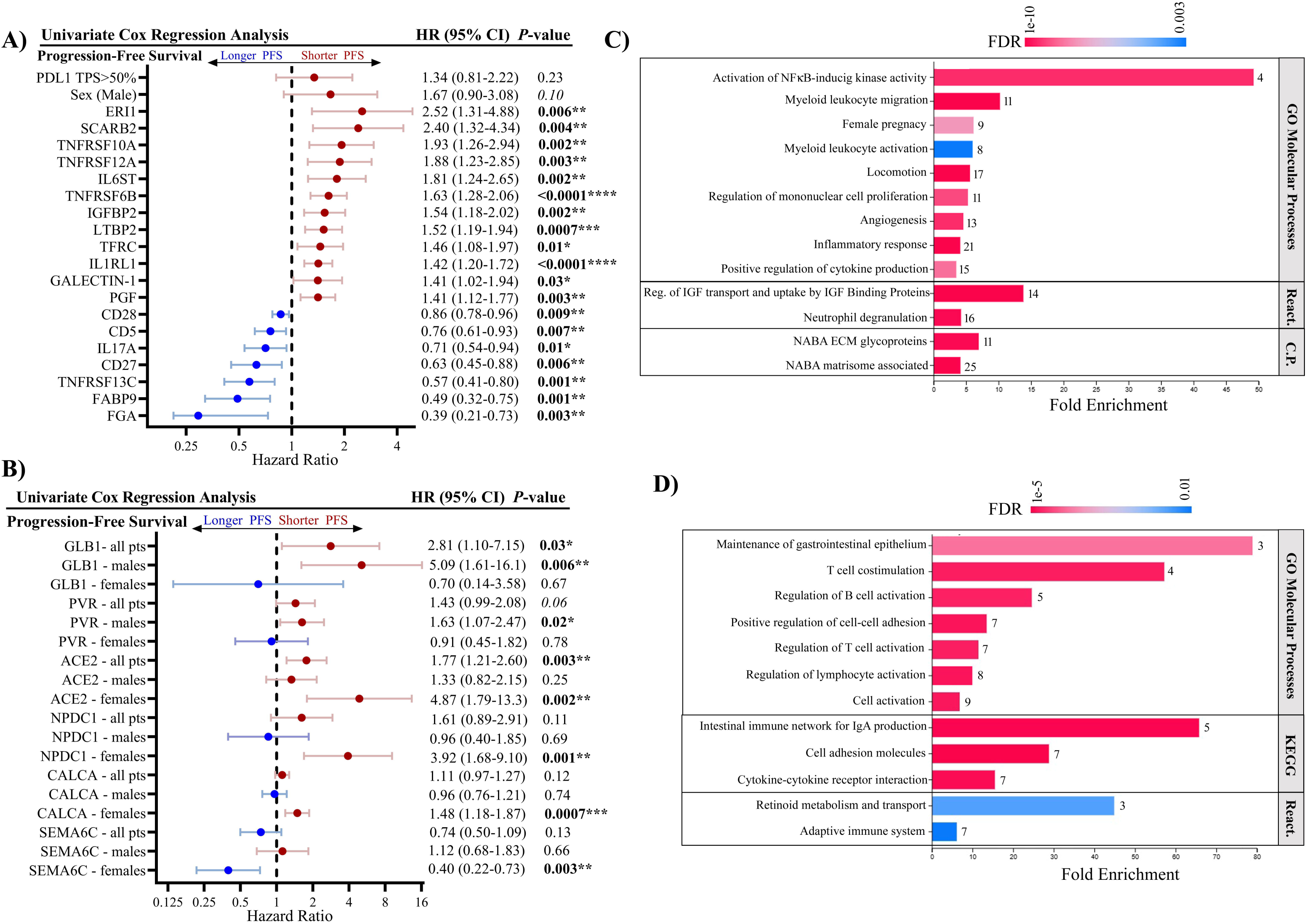
Forest plots of hazard ratios for (A) individual protein biomarkers and (B) sex-related differences in hazard ratios for selected protein biomarkers, Fold enrichment of pathways for proteins significantly associated with (C) shorter PFS (HR>1) or (D) longer PFS (HR<1). Number of proteins enriched in the pathway are indicated to the right of bars. Hazard ratio = HR; PFS = progression-free survival; CI = confidence interval; TPS = Tumour Proportion Score; REACT. = Reactome; C.P. = Canonical Pathways; KEGG = KEGG Pathway; pAdj = adjusted p value. Logrank test (* p<0.05, ** p<0.01, ***p<0.001, ****p<0.0001).

**Figure 4:**
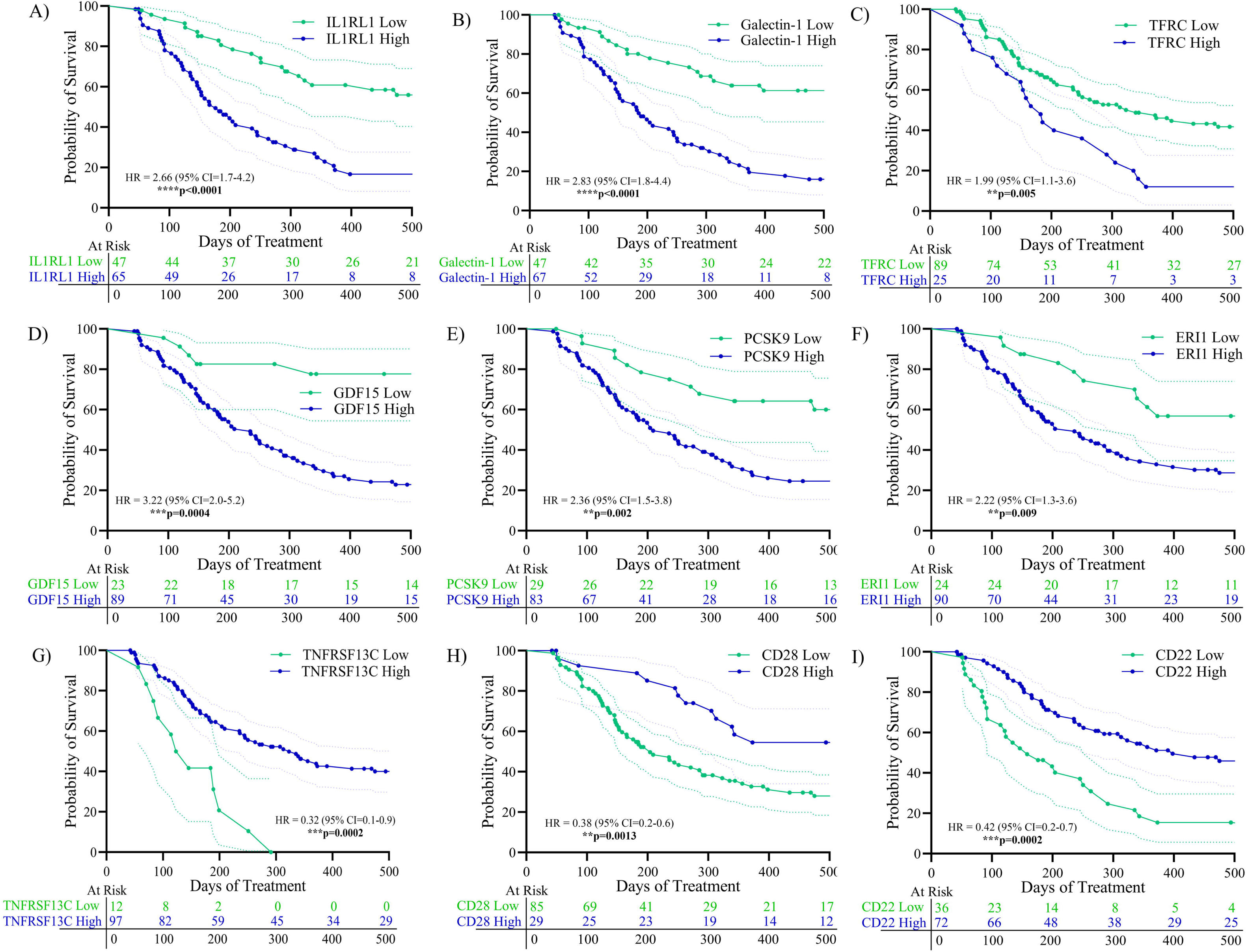
Kaplan–Meier survival curves are shown for patients stratified into high- and low-expression groups for proteins identified as significantly associated with outcome by Cox regression. Proteins with hazard ratio (HR) > 1 (risk-associated) show poorer survival in the high-expression group compared with the low-expression group (A-F), whereas proteins with HR < 1 (protective) show better survival in the high-expression group (G-I). Survival differences between groups were assessed using the log-rank test, with corresponding p values indicated on each plot along with hazard ratio (HR) and confidence intervals. Logrank test (* p<0.05, ** p<0.01, ***p<0.001, ****p<0.0001).

Independent Cox regression analysis within the male and female patient cohorts identified predictive biomarkers with pronounced differences between males and females (Fig. 3B, Supp. Table S2). The senescence marker GLB1 and the immune checkpoint PVR were associated with high risk of progressive disease in males. PVR, is over-expressed by tumours and acts to suppress NK and T cell activity^11^. In the female cohort, ACE2, NPDC1, and the hormone calcitonin (CALCA) were associated with risk of shorter PFS (Fig. 3B).

Proteins that were significantly associated with shorter relapse-free survival (HR>1) were analyzed by pathway enrichment and uncovered over-represented pathways related to NF-κB activation, myeloid leukocyte migration and activation, angiogenesis, ECM remodeling, and neutrophil degranulation (Fig. 3C). Enrichment of these pathways is suggestive of an M2/MDSC dominant TME signature and exclusion of cytotoxic cells via a combination of ECM remodeling and vascular exclusion. Proteins associated with durable response to ICB revealed over-representation of pathways related to activation of T- and B-cells, cell adhesion, and adaptive immune responses that are suggestive of an immune system poised to respond upon release of immune checkpoints by ICI therapy (Fig. 3D). Sex-dependent differences in pathway enrichment were also examined (Supp. Fig. S3).

### Survival Outcomes and Network Analysis Associated with ICI Therapy Response

A subset of the potential prognostic biomarkers was evaluated using K-M survival analysis (Fig. 4). High plasma EV levels of IL1RL1, galectin-1, TFRC, GDF15, PCSK9, and ERI1, were associated with shorter PFS. The IL-33 receptor IL1RL1 was found to be one of the most predictive biomarkers with a mean PFS of 176 days for high IL1RL1 compared to 670 days for low IL1RL1. However, sex-related differences were observed and IL1RL1 was found to be a much better predictor of poor ICI response for males (HR = 4.86, 95% CI 2.11-11.22, p=5×10^-5^) than females (HR = 1.66, 95%CI 0.85-3.27, p=0.14), while TFRC showed stronger predictive power in females (HR = 3.37, 95%CI 1.64-6.94, p=5×10^-4^) compared to males (HR = 1.41, 95% CI 0.67-2.96, p=0.36) (Supp. Fig. S4). Higher EV levels of TNFRSF13C, CD28, and CD22 were associated with significantly longer PFS, thereby supporting their potential prognostic relevance.

STRING analysis was performed to identify functional interactions among proteins significantly associated with patient outcome. A total of 222 proteins significantly associated with either increased risk (red nodes; HR>1) or decreased risk (blue nodes; HR<1) of NDR were included in the analysis and a PPI network was built with a high confidence score (> 0.7) (Fig. 5). STRING analysis demonstrated a highly inter-connected network of proteins, suggesting that our dataset reflects a highly co-ordinated biological programs associated with ICI response (PPI enrichment p-value = 3×10^-15^). Clustering of the network associated with poor ICI response revealed central node centered around the C-X-C motif protein CXCL8, a chemokine associated with tumour neutrophil infiltration and poor response to ICI^12^. Protein networks in blue indicate those associated with durable response and show integrated T cell and B cell networks as well as possible interactions with immune checkpoints and other inhibitory proteins that may moderate responses.

**Figure 5:**
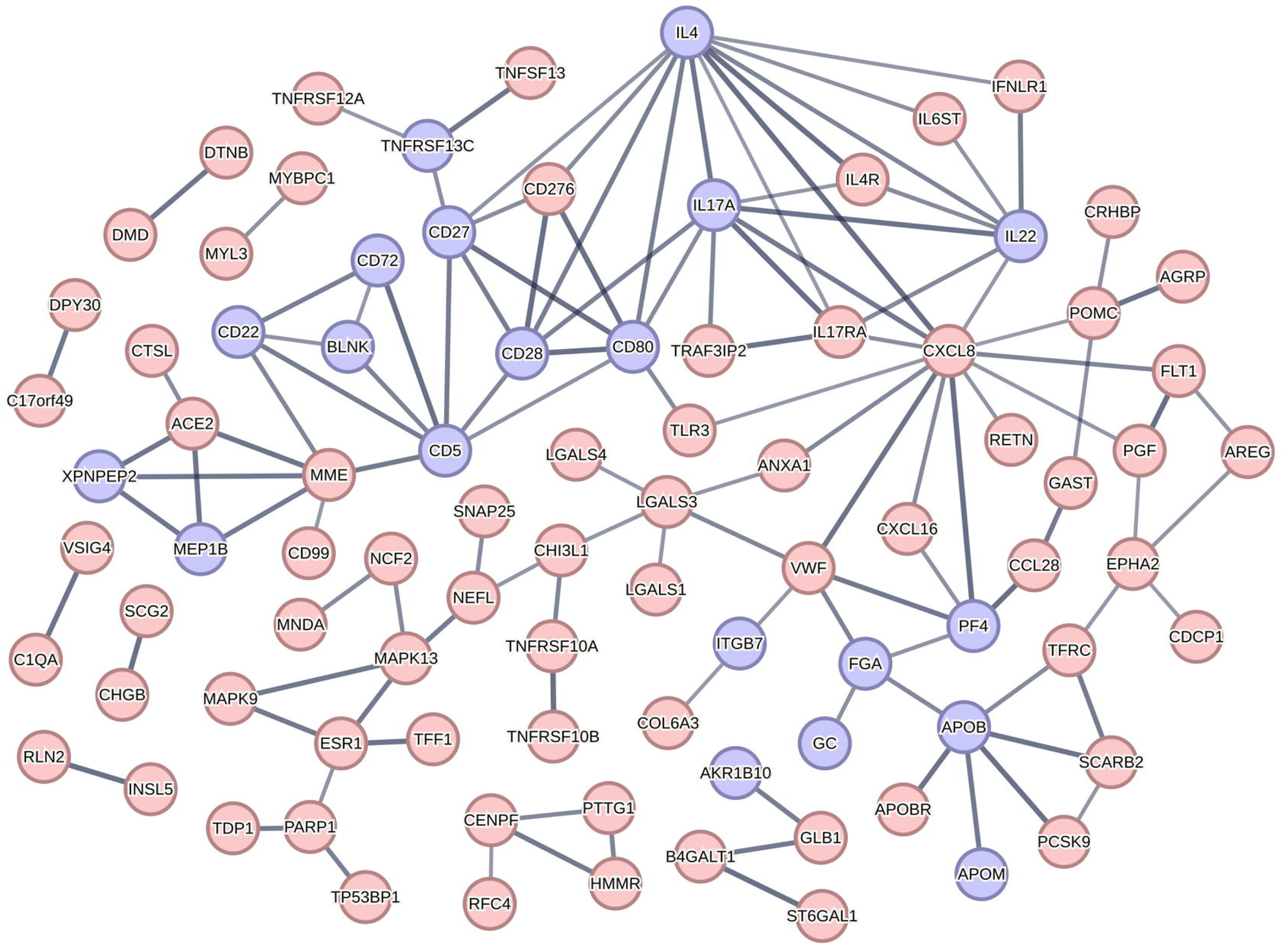
STRING analysis depicting functional interactions among proteins significantly associated with patient outcome. Proteins with hazard ratio (HR) > 1, indicating association with increased risk, are shown as red nodes, while proteins with HR < 1, associated with decreased risk, are shown as blue nodes. Node size reflects protein connectivity, and edge thickness corresponds to interaction confidence scores derived from STRING database analysis. Disconnected nodes are not shown.

### A six-protein EV biomarker signature is predictive of NDR to ICI

Logistic regression was applied to biomarkers that were independently correlated to non-durable response to ICI in our patient cohort and used to generate odds ratios for NDR (Fig. 6A). Twelve candidate biomarkers were identified and combined in a preliminary logistic regression model to discriminate between NDR and DR to ICI treatment and then refined using a backward elimination approach until a six-biomarker model was identified. The model included five biomarkers that were positively correlated to NDR (IL1RL1, TFRC, CCN5, ERI1, and IGFBPL1) and one biomarker, TNFRSF13C, that was negatively correlated to NDR. The good discriminatory performance of the six-biomarker model was confirmed by ROC curve analysis, which showed an improved AUC of 0.907 for the combined model compared to the individual biomarkers (Fig. 6B,C). At the optimal cut-off threshold, a sensitivity of 87.5% (95% CI = 76.8%-94.4%) and a specificity of 83.3% (95% CI = 68.6%-93.0%) was achieved. The model had a positive predictive value of 88.9% (95% CI = 77.8%-95.1%) and a negative predictive value 81.4% (95% CI = 67.5%-92.1%). Based on the probabilities generated by the six-biomarker model, patients were stratified into low risk (p≤0.4), moderate risk (0.4<p<0.6), and high risk (p≥0.6) for NDR. K-M survival analysis demonstrated a clear separation of high and low risk patients based on the probabilities calculated by the biomarker signature (Fig. 6D). High-risk patients had a mean PFS of 158 days, moderate-risk patients had a mean PFS of 739 days, while mean PFS was not reached for low-risk patients. The model was found to have good discriminatory powers for both sexes, with an AUC for males of 0.950 (95% CI = 0.894-1.00) and an AUC of 0.900 (95%CI = 0.806-0.994) for females (Fig. 6E; Suppl. Fig. S5).Overall, these findings indicate that the six-biomarker panel was highly discriminatory for NDR in this exploratory cohort but requires independent validation.

**Figure 6:**
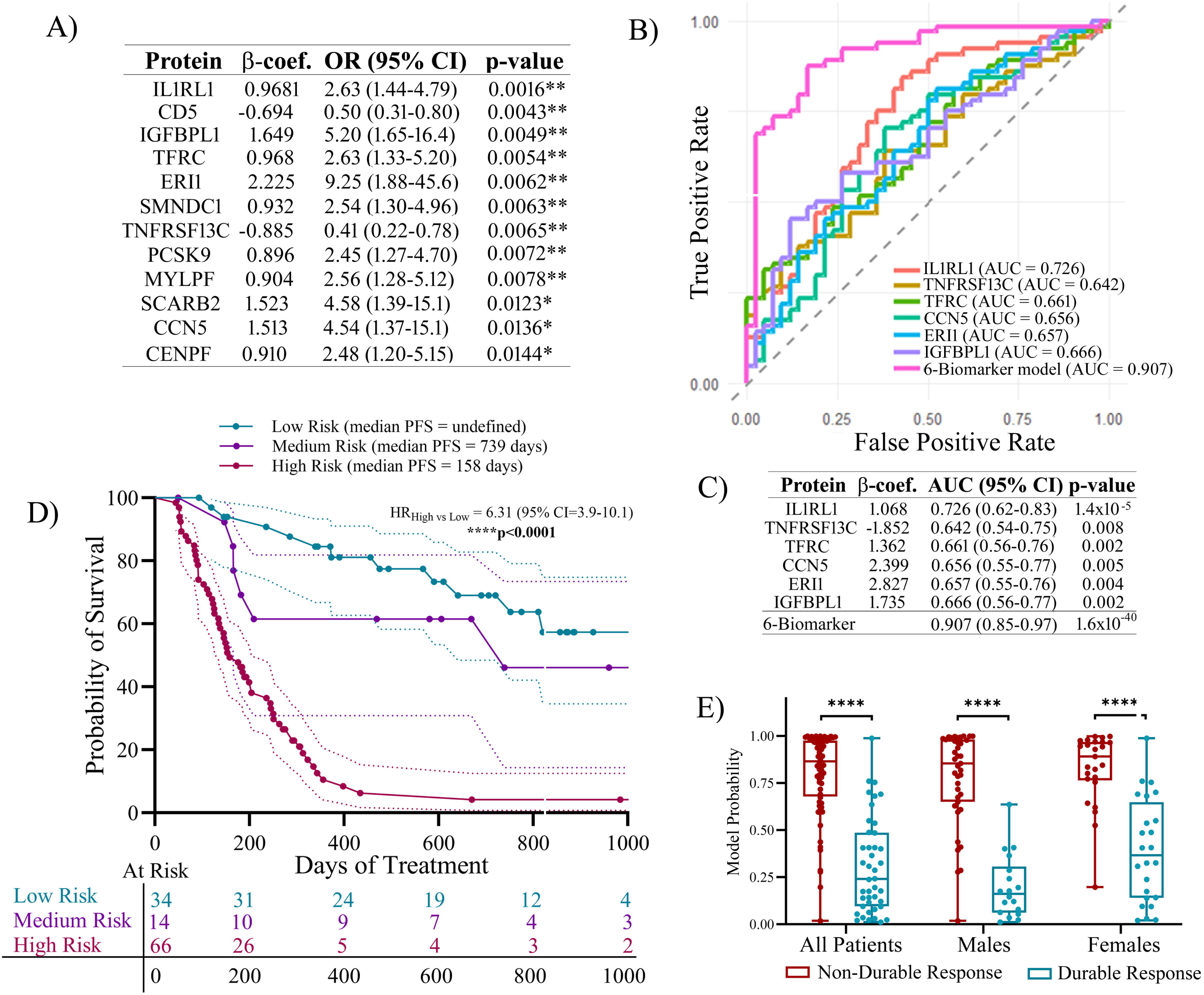
(A) Logistic regression analysis was used to determine the odds ratio (OR) of selected biomarkers for NDR (B) Receiver Operating Characteristic (ROC) curve showing performance of individual and combined biomarkers (6-biomarker model). (C) Area under the curve (AUC) of individual and combined biomarkers. (D) Kaplan-Meier plot showing PFS of patients stratified based on risk as determine by probabilities calculated by the 6-biomarker model. K-M plot of low-risk (p≤0.4), medium-risk (0.4<p<0.6), and high-risk (p≥0.6) patients with logrank test are shown. (E) Boxplot showing stratification of NDR and DR patients based on probabilities calculated by the 6-biomarker model in the entire patient cohort, and males and females separately. β-coef. = β-coefficient; OR = Odds Ratio; AUC = Area Under the Curve; HR = Hazard Ratio; PFS = Progression-free survival. *p<0.05, **p<0.01, ***p<0.001, ****p<0.0001.

## DISCUSSION

The use of therapies targeting immune checkpoints such as PD-1, PD-L1, and CTLA-4 has offered patients suffering from lung and other cancers dramatically improved survival outcomes, but only for a subset of patients. A better understanding of the determinants of adaptive immune responses is required to address treatment resistance and robust predictive biomarkers that can both predict and monitor response are needed to stratify patients who will benefit from ICI treatment from those who would be better served by alternative treatment regimens.

Although PD-L1 tumour expression, tumour mutation burden, and the degree of tumour infiltration with lymphocytes are moderately predictive of ICI response in NSCLC, the use of liquid biopsy-based approaches could yield biomarkers that better reflect disease heterogeneity, tumour–immune interactions and enable longitudinal monitoring of disease response. High throughput plasma proteomics has yielded robust biomarkers for ICI response prediction, including a plasma protein model based on SomaScan profiling of 388 resistance-associated proteins^13^.

Due to the critical role of EVs in mediating the interplay between tumour and immune cells in the TME, influencing immune responses and ultimately the success of ICI therapy, the use of EVs in liquid biopsy biomarker development could have significant clinical implications. Since they are released directly from tumour and immune cells and are enriched in cell-of-origin proteins and immune modulators^4,5^, EVs are a rich resource for biomarker discovery. Although most EV biomarkers published to date have been diagnostic, EV protein profiling has been evaluated as a biomarker strategy for immune checkpoint inhibitor response. These studies have primarily used targeted assessment of selected immune-suppressive molecules such as PD-L1 and TGF-β^5^, low-plex Olink panels with limited predictive power, or mass spectrometry^14^. This study represents the first application of high-plex proteomics for circulating EV-based ICI response prediction.

Many response-associated proteins identified in our study are membrane-associated proteins, making the EV compartment especially biologically relevant. When packaged into EVs, these proteins remain embedded in an organized vesicular membrane, preserving ligand-binding capacity and functional signaling properties^4,5^. Unlike soluble plasma proteins derived from a highly diluted and systemically mixed compartment, selective protein packaging and display on EVs enables them to capture cell-of-origin specific information, including tumour and immune cell identity, activation state, and signaling^4,5^. This is particularly relevant in immuno-oncology, where checkpoint molecules and other immune-modifying surface proteins may be released in vesicular form as part of local and systemic tumour-immune communication^5,15^. EV-associated PD-L1 predicts response to ICB ^15^, remains functionally competent as a ligand capable of engaging PD-1 and suppressing T-cell activity^16^, and strategies to improve ICB response by inhibiting release of PD-L1+ EVs are being explored^17^. Together, these findings support EV-based proteomics as a potentially more mechanistically informative biomarker discovery strategy than bulk plasma, due to the enrichment of tumour- and immune-cell-specific signals^5,11^.

In addition to conventional soluble immune regulatory proteins, our study identified several proteins associated with a high risk of progression on ICI that have been previously implicated in tumour-derived EV-mediated immune modulation. These include members of the TNF superfamily (TNFRSF6B, TNFRSF10A, and TNFRSF12A), galectins (galectin-1 and galectin-3), and immune checkpoint proteins (PD-L1 and CD276). EV-associated galectins have been well characterised as immunosuppressive factors in tumour-derived EVs. Galectin-1 and galectin-3 are packaged into EVs and released from tumour cells into the TME, where they reduce anti-PD-1 efficacy^18^ by suppressing T-cell function^10^, promoting M2 macrophage polarization^19,20^, and activating cancer-associated fibroblasts^18^. TNF superfamily death receptors exposed on EVs have been reported to function as ligand sinks, sequestering cytotoxic ligands such as TRAIL released by CD8⁺ T cells and NK cells. This process reduces effective death receptor signalling and limits immune-mediated tumour cell killing^21,22^. While other TNF superfamily members have been reported on EVs, EV-associated TNFRSF6B is not well established and requires further validation. EV-associated immune checkpoint proteins were also associated with short relapse-free survival in this study, including PD-L1 and CD276, which are known to contribute to NK- and T-cell suppression and immune evasion when present on EVs^11^. Together, these observations support a model in which tumour-derived EVs exposing galectins, TNF receptor family members, and checkpoint proteins provide a potent mechanism of immune evasion that may contribute to the limited durability of response to immune checkpoint inhibition.

The protein interaction network identified in this study revealed a co-ordinated core of highly interacting proteins characteristic of an organized network-level response rather than dysregulation of isolated proteins. In durable responders, a densely connected hub of inter-connected proteins reflecting T-cell and B-cell regulation associated with ICI response was observed, reflecting an immune system primed for response upon release of immune checkpoints. A high risk of progression on ICI was associated with a larger, yet less inter-connected, protein interaction network, suggestive of multiple biological pathways at work rather than a single dominant immune evasion mechanism. This pattern may reflect the heterogeneity of immune evasion mechanisms employed by tumours to evade immune control. In non-durable responders, string analysis revealed a node centered around CXCL8, a CXC chemokine that is secreted by neoplastic cells, including lung cancer^23^, and which mediates metastasis, angiogenesis, drug resistance, and immunosuppressive cell recruitment, particularly neutrophils^24^. Although numerous reports have identified CXCL8 as a predictor of ICI response in NSCLC^12,24,25^ and CXLC8 has been found to be associated with EVs, EV-associated CXCL8 has not previously been reported to be a mediator of ICI response. However, EVs bearing CXCL8 have been reported to promote immune evasion by facilitating transport of CXCL8 into CD8+ T cells to inhibit proliferation by disrupting glucolipid metabolism^26^ and to increase radio-resistance of prostate cancer cells by inducing autophagy^27^.

Several proteins associated with M2 polarization, TAMS, and neutrophils were elevated in NDR, including CH3IL1, IL1RL1, IL6ST (gp130), FGF7, and CD177. EV-associated CD177 has been correlated to emphysema^28^, a risk factor for NSCLC. IL6ST is abundant in tumour-derived EVs where it acts to promote the transformation of macrophages to an immunosuppressive M2 phenotype^29,30^. Other potentially interesting immunosuppressive protein clusters identified were those involved in ECM remodeling (e.g. LTBP2, LTBP3, SPON2, CTSL) and angiogenic–myeloid signaling (PGF, FLT1/VEGFR1, VEGFA, VEGFD), which may lead to leaky vasculature and immune exclusion. Also observed were cytokines, chemokines, and glycoproteins involved in M2 macrophage polarization and MDSC/Treg recruitment (e.g. CXCL8, CCL23, CCL25, GDF15, IL6ST, galectin-1, galectin-3), which work together to support the formation of an immunosuppressive niche.

The influence of sex hormones on cancer pathogenesis and immunotherapy responses is of emerging importance in the field of immuno-oncology. Whereas estrogens contribute to immune evasion via T-cell exhaustion, immune tolerance, and regulation of PD-L1 biology, androgens promote immune suppression by inhibiting immune activation^31^. Estrogen receptor signaling has been implicated in lung cancer development and prognosis following anti-PD-1 therapy^31^ and leads to PD-L1 transcriptional activation via aromatase-induced estradiol production^32^. Therefore, estrogen signaling contributes to an immunosuppressive TME by promoting T cell exclusion and dysfunction and polarization of TAMs towards M2 phenotype^1,31^. The role sex hormones play as key modulators of immune surveillance and ICI responsiveness in NSCLC highlights the importance of incorporating sex-specific analyses into biomarker discovery.

In this study, we report several sex-specific disparities in proteins and pathways associated with response to ICI. In male NDR patients, we observed elevated levels of proteins associated with suppression of T cell responses, including IL1RL1, the immune checkpoints CD276 and PVR, and FNDC1, a protein enriched in tumour stroma that is involved in ECM remodeling and angiogenesis^33^. In females, elevated levels of CEACAM19, a promoter of tumour progression and metastasis^34^, KIR3DL2, an NK-cell immune checkpoint, and TFRC were observed. Although sex-related differences in pathway enrichment were relatively subtle, we observed an enrichment of pathways associated with positive regulation of cytokine production in males, as opposed to over-representation of ECM glycoprotein pathways in females. These findings suggest the intriguing possibility that treatment failure in female patients may be driven by TGF-β/ECM-dominated immune exclusion (TNC, LTBP2, IGFBP2, IGFBP4, MATN2, MATN3), whereas male NDR patients feature angiogenic/myeloid-driven immune exclusion (ANGPT2, PGF, CCL23, CCL28, CXCL16, CTSL). This divergence could help explain variability in ICI responsiveness between sexes and highlights the importance of sex-stratified analyses when identifying biomarkers and designing therapeutic strategies.

A six-biomarker signature that provided good prognostic performance for identifying patients who failed to achieve a durable benefit from ICI treatment was identified in this study. The biomarkers included IL1RL1, TFRC, ERI1, CCN5, IGFBPL1 and TNFSRSF13C. One of the most notable proteins that was increased in NDR was IL1RL1, the only known receptor for IL-33, an alarmin cytokine involved in damage-associated immune responses and tumour-associated Th2-skewed immunity^35^. This receptor is predominantly expressed on Th2-type cells, such as Tregs and TAMs, and the IL-33/ST2 axis has been identified as a potential target to improve immune checkpoint therapy outcomes^36^. TFRC has been identified as a predictor of poor response to anti-PD-1 therapy in HCC^37^. In contrast, ERI1, an RNA processing protein, has not previously been implicated previously in ICI response. Although we did not attempt it in this study due to insufficient patient numbers, incorporating sex-specific biomarker selection can lead to the development of more robust prediction models^38^.

Several of proteins associated with poor response identified in this study have inhibitors that are approved or in clinical development, including PCSK9 and GDF15. PCSK9, best known for its role in LDL receptor regulation, has been implicated in tumour immune evasion via suppression of antigen presentation and T cell responses mediated by reduced recycling of MHC-1 and T cell receptor, respectively^39,40^. GDF15, a stress-response cytokine within the TGF-β superfamily, is over-expressed in multiple solid tumours, including lung, and contributes to the establishment of an immune-suppressive TME^41^. Circulating levels of PCSK9 and GDF15 have been reported to be associated with outcomes following anti-PD-1 therapy in NSCLC^42,43^. PCSK9 blockade has been shown to enhance PD-1 efficacy by increasing intra-tumoural CD8^+^ T-cell infiltration and reducing Treg accumulation^39,44^, while GDF15 targeting reprograms the immunosuppressive tumor microenvironment and amplifies anti-PD-1 efficacy in vivo, including in NSCLC models^45^. Collectively, these findings support large-scale EV proteomics as a strategy to identify biologically relevant pathways associated with immune resistance and therapeutic targets with the potential to improve responses to PD-1/PD-L1 inhibition.

Limitations of the current study include its exploratory nature, which limits the strength of inferences that can be made regarding the identified biomarkers and putative evasion mechanisms; further independent studies are therefore required to substantiate our findings. Furthermore, plasma EV-based studies can be difficult to reproduce across sites, owing to differences in EV isolation methodologies and plasma preparation protocols. Proteomic analyses are also strongly influenced by the methodology used. While PEA assays show good inter-assay correlation with one another, they have been reported to correlate poorly with other proteomic approaches, potentially due to poor overlap in protein coverage across technologies^46^.

Our study demonstrates the feasibility of using PEA-based proteomics to profile baseline circulating EVs for biomarker discovery in cancer immuno-oncology. We propose that interrogating circulating EVs for biomarkers associated with response to ICB may provide a “biological snapshot” of tumour-immune interactions and underlying immune evasion pathways that contribute to treatment resistance. Independent validation studies to confirm these findings are currently ongoing.

## Data Availability

All data produced in the present study are available upon reasonable request to the authors

## Data Availability Statement

The dataset used in this study is available as a supplemental data file.

## Acknowledgments

The authors would like to acknowledge the contribution and continued support of the CHU Dumont Biobank as well as the contribution of Robert Cormier and Alexandre-James Roussel from the ACRI sequencing facility for their NGS expertise. Financial support for this work was provided by: the Canadian Cancer Society through an Atlantic Cancer Research Grant (ACR-21), the David and Nancy Holt Partnership Fund, Terry Fox Research Institute-Marathon of Hope Cancer Center (ACC), New Brunswick Innovation Foundation (NBIF) – Research Professionals Initiative, NBIF – Equipment Grant, and the RR Leger-New Brunswick Health Research Chair to RJO.

## Author contributions

C.T.: Conceptualization, Investigation, Methodology, Data Curation, Formal Analysis, Visualization, Writing – original draft, Writing -review & editing. M.D.: Methodology, Investigation. E.A.: Formal Analysis. A.S.C.: Formal analysis. N.C.: Methodology (Olink automation and NGS). N.F.: Resources. M.A.: Resources. R.J.O.: Conceptualization, Funding, Project Administration, Resources, Supervision, Writing -review & editing.

## References

1. Conforti F, Pala L, Di Mitri D, et al. Sex hormones, the anticancer immune response, and therapeutic opportunities. Cancer Cell 2025; 43(3):343–360. doi:10.1016/j.ccell.2025.02.013

2. Carbone DP, Reck M, Paz-Ares L, et al. First-Line Nivolumab in Stage IV or Recurrent Non-Small-Cell Lung Cancer. N Engl J Med 2017; 376(25):2415–2426. doi:10.1056/NEJMoa161349

3. Zhou S, Yang H. Immunotherapy resistance in non-small-cell lung cancer: From mechanism to clinical strategies. Front Immunol 2023; 14:1129465. doi:10.3389/fimmu.2023.1129465

4. Kalluri R. The biology and function of extracellular vesicles in immune response and immunity. Immunity 2024; 57(8):1752–1768. doi: 10.1016/j.immuni.2024.07.009.

5. Asleh K, Dery V, Taylor C, et al. Extracellular vesicle-based liquid biopsy biomarkers and their application in precision immuno-oncology. Biomark Res 2023;11(1):99. doi:10.1186/s40364-023-00540-2

6. Akbar S, Raza A, Mohsin R, et al. Circulating exosomal immuno-oncological checkpoints and cytokines are potential biomarkers to monitor tumor response to anti-PD-1/PD-L1 therapy in non-small cell lung cancer patients. Front Immunol 2023; 13:1097117 doi:10.3389/fimmu.2022.1097117

7. Knol JC, de Reus I, Schelfhorst T, et al. Peptide-mediated ’miniprep’ isolation of extracellular vesicles is suitable for high-throughput proteomics. EuPA Open Proteom 2016; 11:11–15. doi:10.1016/j.euprot.2016.02.001

8. Szklarczyk D, Gable AL, Lyon D, et al. STRING v11: protein-protein association networks with increased coverage, supporting functional discovery in genome-wide experimental datasets. Nucleic Acids Res 2019; 47(D1):D607–D613. doi:10.1093/nar/gky1131

9. Lu Y, Huang P, Zeng X, et al. Inhibition of FNDC1 suppresses gastric cancer progression by interfering with Gβγ-VEGFR2 complex formation. iScience 2023; 26(9):107534. doi:10.1016/j.isci.2023.107534

10. Maybruck BT, Pfannenstiel LW, Diaz-Montero M, Gastman BR. Tumor-derived exosomes induce CD8^+^ T cell suppressors. J Immunother Cancer 2017; 5(1):65. doi:10.1186/s40425-017-0269-7

11. Jeppesen DK, Sanchez ZC, Kelley NM, et al. Blebbisomes are large, organelle-rich extracellular vesicles with cell-like properties. Nat Cell Biol 2025; 27(3):438–448. doi:10.1038/s41556-025-01621-0

12. Schalper KA, Carleton M, Zhou M, et al. Elevated serum interleukin-8 is associated with enhanced intratumor neutrophils and reduced clinical benefit of immune-checkpoint inhibitors. Nat Med 2020; 26(5):688–692. doi:10.1038/s41591-020-0856-x

13. Yellin B, Lahav C, Sela I, et al. Analytical validation of the PROphet test for treatment decision-making guidance in metastatic non-small cell lung cancer. J Pharm Biomed Anal 2024; 238:115803. doi: 10.1016/j.jpba.2023.115803.

14. Trilla-Fuertes L, Gámez-Pozo A, Laso-García F, et al. Protein content of extracellular vesicles from patients with advanced melanoma changes upon progression to anti-PD1 therapy. Sci Rep 2026; 16(1):5891. doi:10.1038/s41598-026-35848-0

15. Kumar, M.A., Baba, S.K., Sadida, H.Q. et al. Extracellular vesicles as tools and targets in therapy for diseases. Sig Transduct Target Ther 2024; 9(1):27 (2024). doi: 10.1038/s41392-024-01735-1.

16. Chen G, Huang AC, Zhang W, et al. Exosomal PD-L1 contributes to immunosuppression and is associated with anti-PD-1 response. Nature 2018; 560(7718):382–386. doi:10.1038/s41586-018-0392-8

17. Wu B, Huang X, Shi X, Jiang M, Liu H, Zhao L. LAMTOR1 decreased exosomal PD-L1 to enhance immunotherapy efficacy in non-small cell lung cancer. Mol Cancer 2024; 23(1):184. doi:10.1186/s12943-024-02099-4

18. Fan Y, Song S, Pizzi MP, et al. Exosomal Galectin-3 promotes peritoneal metastases in gastric adenocarcinoma via microenvironment alterations. iScience 2024; 28(1):111564. doi:10.1016/j.isci.2024.111564

19. Li J, Pan Y, Yang J, et al. Tumor necrosis factor-α-primed mesenchymal stem cell-derived exosomes promote M2 macrophage polarization *via* Galectin-1 and modify intrauterine adhesion on a novel murine model. Front Immunol 2022; 13:945234. doi:10.3389/fimmu.2022.945234

20. Wang M, Sun Y, Gu R, et al. Shikonin reduces M2 macrophage population in ovarian cancer by repressing exosome production and the exosomal galectin 3-mediated β-catenin activation. J Ovarian Res 2024; 17(1):101. doi:10.1186/s13048-024-01430-3

21. Setroikromo R, Zhang B, Reis CR, et al. Death Receptor 5 Displayed on Extracellular Vesicles Decreases TRAIL Sensitivity of Colon Cancer Cells. Front Cell Dev Biol 2020; 8:318. doi:10.3389/fcell.2020.00318

22. Canas JJ, Enslow SM, Bhimani S, et al. Extracellular vesicles decoying across host immunity. J Leukoc Biol. 2025; 118(1):qiaf173. doi:10.1093/jleuko/qiaf173

23. Ma X, Zhu X, Zou M, et al. Expression of CXCL8 and its relationship with prognosis in patients with non-small cell lung cancer. Am J Cancer Res. 2024; 14(6):2934–2945. doi: 10.62347/LJDQ3897

24. Meier C, Brieger A. The role of IL-8 in cancer development and its impact on immunotherapy resistance. Eur J Cancer 2025; 218:115267. doi: 10.1016/j.ejca.2025.115267.

25. Belluomini L, Cesta Incani U, Smimmo A, et al. Prognostic impact of Interleukin-8 levels in lung cancer: A meta-analysis and a bioinformatic validation. Lung Cancer 2024; 194:107893. doi:10.1016/j.lungcan.2024.107893

26. Xu F, Wang X, Huang Y, et al. Prostate cancer cell-derived exosomal IL-8 fosters immune evasion by disturbing glucolipid metabolism of CD8^+^ T cell. Cell Rep 2023; 42(11):113424. doi: 10.1016/j.celrep.2023.113424.

27. Wang, X., Xu, F., Kou, H., et al. (2023). Stromal cell-derived small extracellular vesicles enhance radioresistance of prostate cancer cells via interleukin-8-induced autophagy. J Extracell Vesicles 2023;12(7):e12342. doi:10.1002/jev2.12342

28. Koba T, Takeda Y, Narumi R, et al. Proteomics of serum extracellular vesicles identifies a novel COPD biomarker, fibulin-3 from elastic fibres. ERJ Open Res 2021; 7: 00658–2020. doi: 10.1183/23120541.00658-2020

29. Ling HY, Yang Z, Wang PJ, et al. Diffuse large B-cell lymphoma-derived exosomes push macrophage polarization toward M2 phenotype via GP130/STAT3 signaling pathway. Chem Biol Interact 2022; 352:109779. doi:10.1016/j.cbi.2021.109779

30. Ham S, Lima LG, Chai EPZ, et al. Breast Cancer-Derived Exosomes Alter Macrophage Polarization *via* gp130/STAT3 Signaling. Front Immunol 2018;9:871. doi:10.3389/fimmu.2018.00871

31. Rodriguez-Lara V, Soca-Chafre G, Avila-Costa MR, et al. Role of sex and sex hormones in PD-L1 expression in NSCLC: clinical and therapeutic implications. Front Oncol 2023; 13:1210297. doi: 10.3389/fonc.2023.1210297

32. Anobile DP, Salaroglio IC, Tabbò F, et al. Autocrine 17-β-Estradiol/Estrogen Receptor-α Loop Determines the Response to Immune Checkpoint Inhibitors in Non-Small Cell Lung Cancer. Clin Cancer Res 2023; 29(19):3958–3973. doi:10.1158/1078-0432.CCR-22-3949

33. Lu Y, Huang P, Zeng X, et al. Inhibition of FNDC1 suppresses gastric cancer progression by interfering with Gβγ-VEGFR2 complex formation. iScience 2023; 26(9):107534. doi:10.1016/j.isci.2023.107534

34. Beauchemin N, Arabzadeh A. Carcinoembryonic antigen-related cell adhesion molecules (CEACAMs) in cancer progression and metastasis. Cancer Metastasis Rev 2013; 32:643–671. doi:10.1007/s10555-013-9444-6

35. Schmitz J., Owyang A., Oldham E., et al. IL-33, an interleukin-1-like cytokine that signals via the IL-1 receptor-related protein ST2 and induces T helper type 2-associated cytokines. Immunity 2005; 23:479–490. doi: 10.1016/j.immuni.2005.09.015.

36. Jovanovic M, Gajovic N, Jocic M, et al. Dual targeting of PD-1/PD-L1 and iL-33/ST2 signalling pathways: a promising approach in breast cancer immunotherapy. Ann Med. 2025; 57(1):2593198. doi: 10.1080/07853890.2025.2593198

37. Song F, Hu B, Liang XL, et al. Anlotinib potentiates anti-PD1 immunotherapy via transferrin receptor-dependent CD8^+^ T-cell infiltration in hepatocellular carcinoma. Clin Transl Med 2024; 14(8):e1738. doi:10.1002/ctm2.1738

38. Xie R, Vlaski T, Sha S, et al. Sex-specific proteomic signatures improve cardiovascular risk prediction for the general population without cardiovascular disease or diabetes. J Adv Res 2026; 79:599–606. doi:10.1016/j.jare.2025.03.034

39. Liu X, Bao X, Hu M, et al. Inhibition of PCSK9 potentiates immune checkpoint therapy for cancer. Nature 2020;588(7839):693–698. doi: 10.1038/s41586-020-2911-7

40. Yuan J, Cai T, Zheng X, et al. Potentiating CD8+ T cell antitumor activity by inhibiting PCSK9 to promote LDLR-mediated TCR recycling and signaling. Protein Cell 2021; 12(4):240–260. doi: 10.1007/s13238-021-00821-2

41. Zhang P, Liu Y, Yang H, et al. Inhibition of GDF15/GFRAL: A novel opportunity for the treatment of solid tumors. Int Immunopharmacol 2026; 170:116109. doi: 10.1016/j.intimp.2025.116109

42. Xie M, Yu X, Chu X, et al. Low baseline plasma PCSK9 level is associated with good clinical outcomes of immune checkpoint inhibitors in advanced non-small cell lung cancer. Thorac Cancer 2022; 13(3):353–360. doi: 10.1111/1759-7714.14259.

43. Hong G, Sun P, Chung C, et al. Plasma GDF15 levels associated with circulating immune cells predict the efficacy of PD-1/PD-L1 inhibitor treatment and prognosis in patients with advanced non-small cell lung cancer. J Cancer Res Clin Oncol 2023; 149:159–171. doi: 10.1007/s00432-022-04500-5

44. Wang R, Liu H, He P, et al. Inhibition of PCSK9 enhances the antitumor effect of PD-1 inhibitor in colorectal cancer by promoting the infiltration of CD8+ T cells and the exclusion of Treg cells. Front Immunol 2022; 13:947756. doi: 10.3389/fimmu.2022.947756

45. Zhang X, Yin Z, Chen X, et al. Machine learning–guided single-cell multiomics uncovers GDF15-driven immunosuppressive niches in NSCLC: a translational framework for overcoming anti-PD-1 resistance. Transl Oncol 2025; 59:102459. doi: 10.1016/j.tranon.2025.102459

46. Kirsher DY, Chand S, Phong A, et al. Current landscape of plasma proteomics from technical innovations to biological insights and biomarker discovery. Commun Chem 2025; 8(1):279. doi:10.1038/s42004-025-01665-1

