## Supplemental Data for "Immune Checkpoint Response Profiles and Resistance Mechanisms in NSCLC Revealed by Circulating Extracellular Vesicle Proteomics"

### Supplemental Tables

Supplemental Table S1: A comparison of univariate (UVA) and multivariate (MVA; adjusted for age and sex) cox regression analysis showing hazard ratios, confidence intervals, logrank p values, and false discovery rates (FDR) for selected protein biomarkers.

Supplemental Table S2: A comparison of univariate (UVA) cox regression analysis between males and females showing hazard ratios, confidence intervals, logrank p values, and false discovery rates for selected protein biomarkers.

### Supplemental Figures

Supplemental Figure S1: Principal component analysis (PCA) plots of showing clustering of assays (A) before and (B) after removal of outlier assays from the analysis.

Supplemental Figure S2: Boxplots showing FNDC1 protein levels in (A) patients who did or did not experience durable response (B) or between males and females who did or did not experience durable response. Resp. = Response. Mann-Whitney U test (*p<0.05, **p<0.01, ***p<0.001, ****p<0.0001).

Supplemental Figure S3: Fold enrichment of pathways for proteins significantly associated with shorter PFS (HR>1) for either males or females. Number of proteins enriched in the pathway are indicated to the right of bars. G.M. = GO Molecular Processes; R. = Reactome; C.P. = Canonical Pathways; pAdj = adjusted p value.

Supplemental Figure S4: Kaplan–Meier survival curves are shown for both male and female patients stratified into high- and low-expression groups for proteins identified as significantly associated with outcome by Cox regression. Survival differences between groups were assessed using the log-rank test, with corresponding P values indicated on each plot along with hazard ration (HR) and confidence intervals. Logrank test (* $p<0.05$, ** $p<0.01$, ***$p<0.001$, ****$p<0.0001$).

Supplemental Figure S5: Receiver Operating Characteristic (ROC) curve showing performance of six-biomarker model in (A) all patients, and either (B) males or (C) females separately. (D) Table showing area under the curve (AUC) and respective p-value derived from the ROC curves as well as sensitivity and specificity of the six-biomarker model in the entire patient cohort, and males and females separately.

**Supp. Table S1**

| | Univariate Cox Analysis | | | Multivariate Cox Analysis | | |
|---|---|---|---|---|---|---|
| **Protein** | **UVA** | **p-value** | **FDR** | **MVA** | **p-value** | **FDR** |
| IL1RL1 | 1.43 (1.28-2.06) | 0.00007 | 0.113 | 1.36 (1.14-1.64) | 0.0009 | 0.197 |
| FNDC1 | 1.15 (1.07-1.24) | 0.0002 | 0.172 | 1.16 (1.08-1.25) | 0.00006 | 0.057 |
| FLT1 (VEGFR1) | 1.44 (1.04-1.98) | 0.026 | 0.571 | 1.533 (1.10-2.14) | 0.011 | 0.446 |
| VEGFA | 1.15 (1.00-1.33) | 0.05 | 0.653 | 1.19 (1.03-1.39) | 0.01` | 0.523 |
| PGF (PIGF) | 1.41 (1.12-1.77) | 0.003 | 0.317 | 1.48 (1.17-1.86) | 0.0009 | 0.197 |
| Galectin 1 | 1.41 (1.02-1.94) | 0.035 | 0.620 | 1.41 (1.02-1.94) | 0.036 | 0.549 |
| Galectin 3 | 1.62 (1.03-2.56) | 0.038 | 0.633 | 1.56 (0.98-2.49) | 0.061 | 0.605 |
| Galectin 4 | 1.35 (1.00-1.82) | 0.047 | 0.642 | 1.40 (1.02-1.91) | 0.034 | 0.549 |
| TNFRSF6B | 1.62 (1.28-2.06) | 0.00007 | 0.113 | 1.59 (1.25-2.03) | 0.0002 | 0.098 |
| TNFRSF10A | 1.92 (1.26-2.85) | 0.002 | 0.287 | 2.02 (1.33-3.07) | 0.0009 | 0.196 |
| TNFRSF10B | 1.49 (1.00-2.22) | 0.048 | 0.648 | 1.45 (0.97-2.19) | 0.073 | 0.605 |
| TNFRSF12A | 1.88 (1.23-2.85) | 0.003 | 0.317 | 2.06 (1.33-3.18) | 0.0009 | 0.219 |
| LTBP2 | 1.52 (1.19-1.94) | 0.0007 | 0.228 | 1.54 (1.20-1.97) | 0.0009 | 0.197 |
| TNFRSF13C | 0.57 (0.41-0.80) | 0.0009 | 0.258 | 0.62 (0.44-0.85) | 0.004 | 0.316 |
| ARHGEF5 | 1.59 (1.19-1.94) | 0.0005 | 0.228 | 1.78 (1.34-2.35) | 0.00006 | 0.057 |
| SPON2 | 1.71 (1.27-2.30) | 0.0004 | 0.228 | 1.90 (1.38-2.58) | 0.00006 | 0.057 |
| XPNPEP2 | 0.69 (0.56-0.84) | 0.0004 | 0.228 | 0.71 (0.57-0.87) | 0.0008 | 0.197 |
| MAPK9 | 1.91 (1.31-2.78) | 0.0007 | 0.228 | 1.87 (1.27-2.76) | 0.001 | 0.238 |
| SMNDC1 | 1.42 (1.16-1.76) | 0.0008 | 0.228 | 1.48 (1.20-1.83) | 0.0002 | 0.106 |
| IL6ST | 1.81 (1.24-2.64) | 0.002 | 0.288 | 1.81 (1.21-2.71) | 0.004 | 0.316 |
| CXCL8 (IL-8) | 1.26 (1.00-1.57) | 0.045 | 0.642 | 1.30 (1.04-1.61) | 0.021 | 0.523 |
| PD-L1 | 1.13 (1.00-1.29) | 0.056 | 0.655 | 1.15 (1.00-1.32) | 0.047 | 0.574 |

**Supp. Table S2**

| | Males | | | Females | | |
|---|---|---|---|---|---|---|
| **Protein** | **UVA** | **p-value** | **FDR** | **MVA** | **p-value** | **FDR** |
| TNFRSF13C | 0.51 (0.35-0.75) | 0.0006 | 0.57 | 0.79 (0.43-1.47) | 0.46 | 0.99 |
| FNDC1 | 1.20 (1.08-1.34) | 0.0008 | 0.57 | 1.11 (0.10-1.23) | 0.06 | 0.99 |
| SAP18 | 0.49 (0.32-0.75) | 0.0009 | 0.57 | 1.12 (0.89-1.39) | 0.32 | 0.99 |
| TNFRSF6B | 1.58 (1.20-2.07) | 0.001 | 0.57 | 1.86 (1.15-3.00) | 0.011 | 0.97 |
| NEFL | 1.25 (1.09-1.42) | 0.001 | 0.57 | 0.93 (0.79-1.10) | 0.41 | 0.99 |
| TMEM132A | 1.40 (1.14-1.71) | 0.001 | 0.57 | 1.16 (0.93-1.46) | 0.19 | 0.99 |
| HRG | 0.18 (0.06-0.55) | 0.002 | 0.63 | 0.83 (0.42-1.65) | 0.61 | 0.99 |
| CBS | 2.16 (1.31-3.55) | 0.002 | 0.63 | 1.18 (0.78-1.78) | 0.42 | 0.99 |
| SPON2 | 1.86 (1.24-2.78) | 0.002 | 0.63 | 2.02 (1.21-3.38) | 0.007 | 0.84 |
| IL1RL1 | 1.34 (1.08-1.66) | 0.008 | 0.77 | 1.51 (1.05-2.18) | 0.02 | 0.99 |
| GAGE2A | 1.53 (1.07-2.25) | 0.02 | 0.90 | 0.76 (0.64-0.89) | 0.0007 | 0.72 |
| ANGPT2 | 1.72 (1.13-2.60) | 0.01 | 0.81 | 1.16 (0.78-1.73) | 0.46 | 0.99 |
| CTSL | 1.82 (1.08-3.09) | 0.02 | 0.90 | 1.86 (1.07-3.24) | 0.03 | 0.99 |
| CKB | 1.22 (0.86-1.74) | 0.26 | 0.90 | 2.47 (1.44-1.87) | 0.0008 | 0.72 |
| SCN4B | 1.06 (0.95-1.18) | 0.29 | 0.90 | 1.21 (1.08-1.36) | 0.001 | 0.72 |
| GDF15 | 1.20 (0.90-1.61) | 0.22 | 0.90 | 1.94 (1.27-2.97) | 0.002 | 0.72 |
| STAU1 | 1.40 (0.94-2.10) | 0.10 | 0.90 | 2.41 (1.34-4.32) | 0.003 | 0.72 |
| IGFBP2 | 1.33 (0.95-1.86) | 0.10 | 0.90 | 1.96 (1.25-3.06) | 0.003 | 0.72 |
| LTBP2 | 1.47 (1.10-1.97) | 0.01 | 0.77 | 1.86 (1.19-2.90) | 0.006 | 0.82 |
| FLT1 (VEGFR1) | 1.22 (0.79-1.90) | 0.36 | 0.90 | 1.88 (1.22-2.89) | 0.004 | 0.72 |
| KIR3DL2 | 1.24 (0.86-1.79) | 0.26 | 0..90 | 1.92 (1.20-3.1) | 0.007 | 0.84 |
| ERI1 | 1.94 (0.83-4.55) | 0.13 | 0.90 | 3.81 (1.26-11.5) | 0.02 | 0.97 |
| TFRC | 1.60 (0.97-2.64) | 0.07 | 0.90 | 1.59 (1.05-2.41) | 0.03 | 0.99 |
| TNC | 1.22 (0.77-1.95) | 0.38 | 0.90 | 1.87 (1.01-3.45) | 0.04 | 0.99 |
| CCL23 | 1.45 (1.01-2.08) | 0.04 | 0.90 | 1.51 (1.01-2.26) | 0.05 | 0.99 |
| CCL28 | 1.61 (1.09-2.41) | 0.02 | 0.90 | 1.19 (0.83-1.72) | 0.35 | 0.99 |

**Supp. Fig. S1**

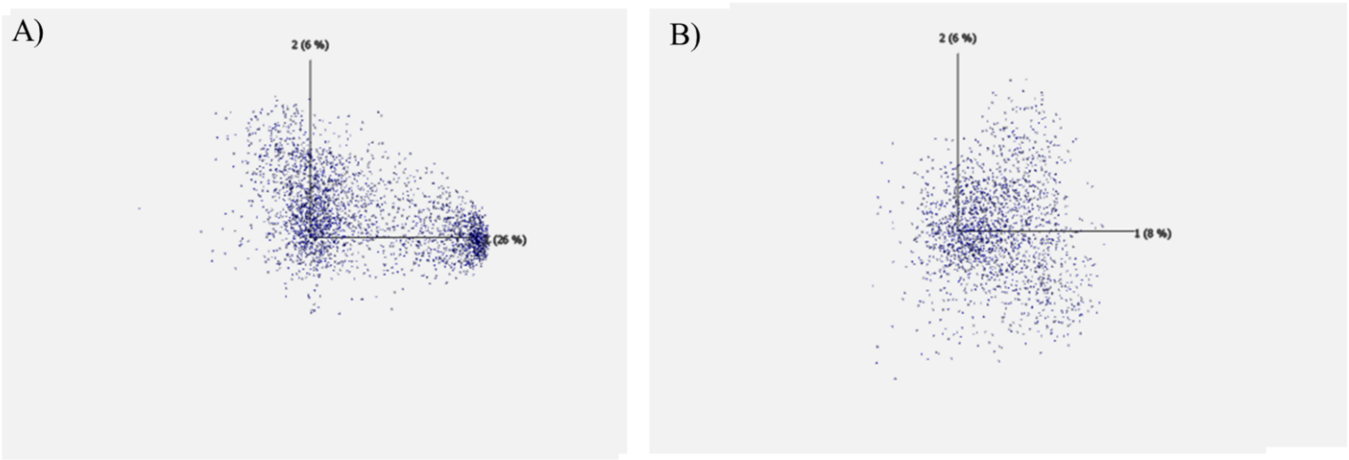


**Supp. Fig. S2**

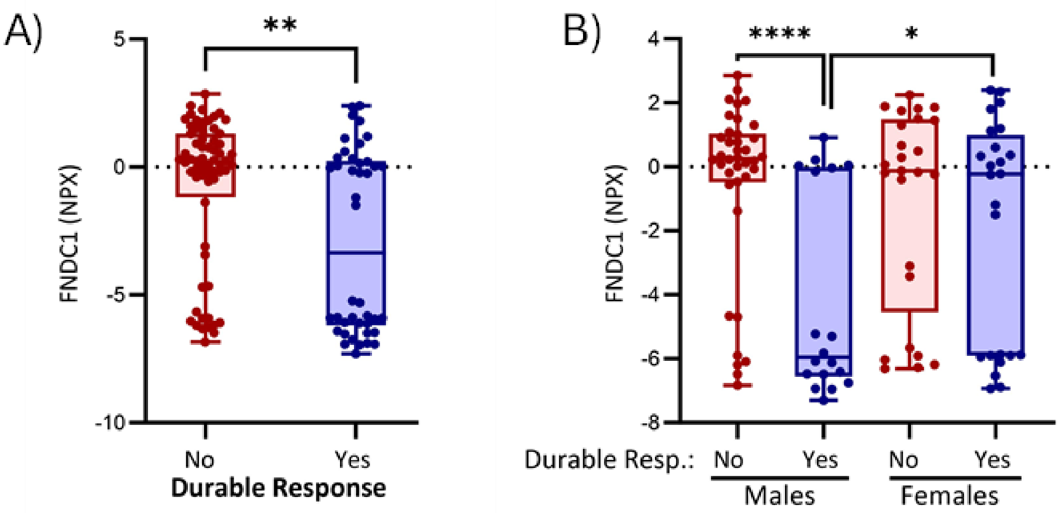

**Supp. Fig. S3**

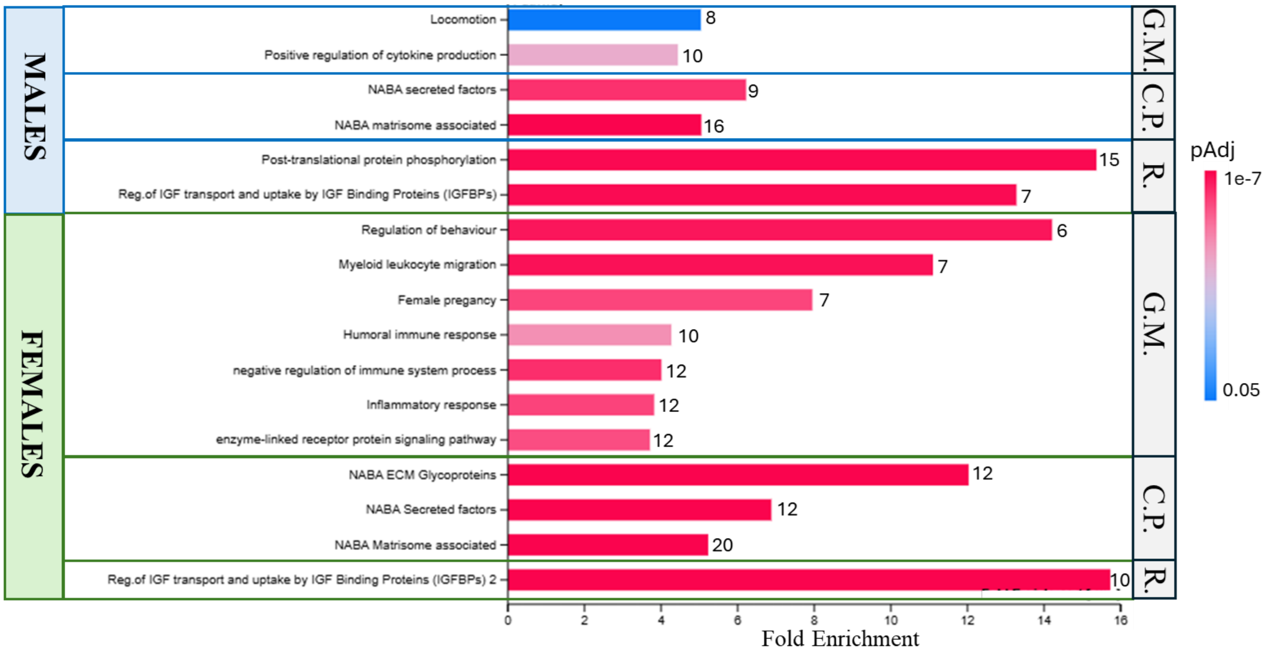

MALES
FEMALES
Locomotion
Positive regulation of cytokine production
NABA secreted factors
NABA matrisome associated
Post-translational protein phosphorylation
Reg.of IGF transport and uptake by IGF Binding Proteins (IGFBPs)
Regulation of behaviour
Myeloid leukocyte migration
Female pregancy
Humoral immune response
negative regulation of immune system process
Inflammatory response
enzyme-linked receptor protein signaling pathway
NABA ECM Glycoproteins
NABA Secreted factors
NABA Matrisome associated
Reg.of IGF transport and uptake by IGF Binding Proteins (IGFBPs) 2
8
10
9
16
15
7
6
7
7
10
12
12
12
12
12
20
10
G.M.
C.P.
R.
G.M.
C.P.
R.
pAdj
1e-7
0.05
0
2
4
6
8
10
12
14
16
Fold Enrichment

**Supp. Fig. S4**

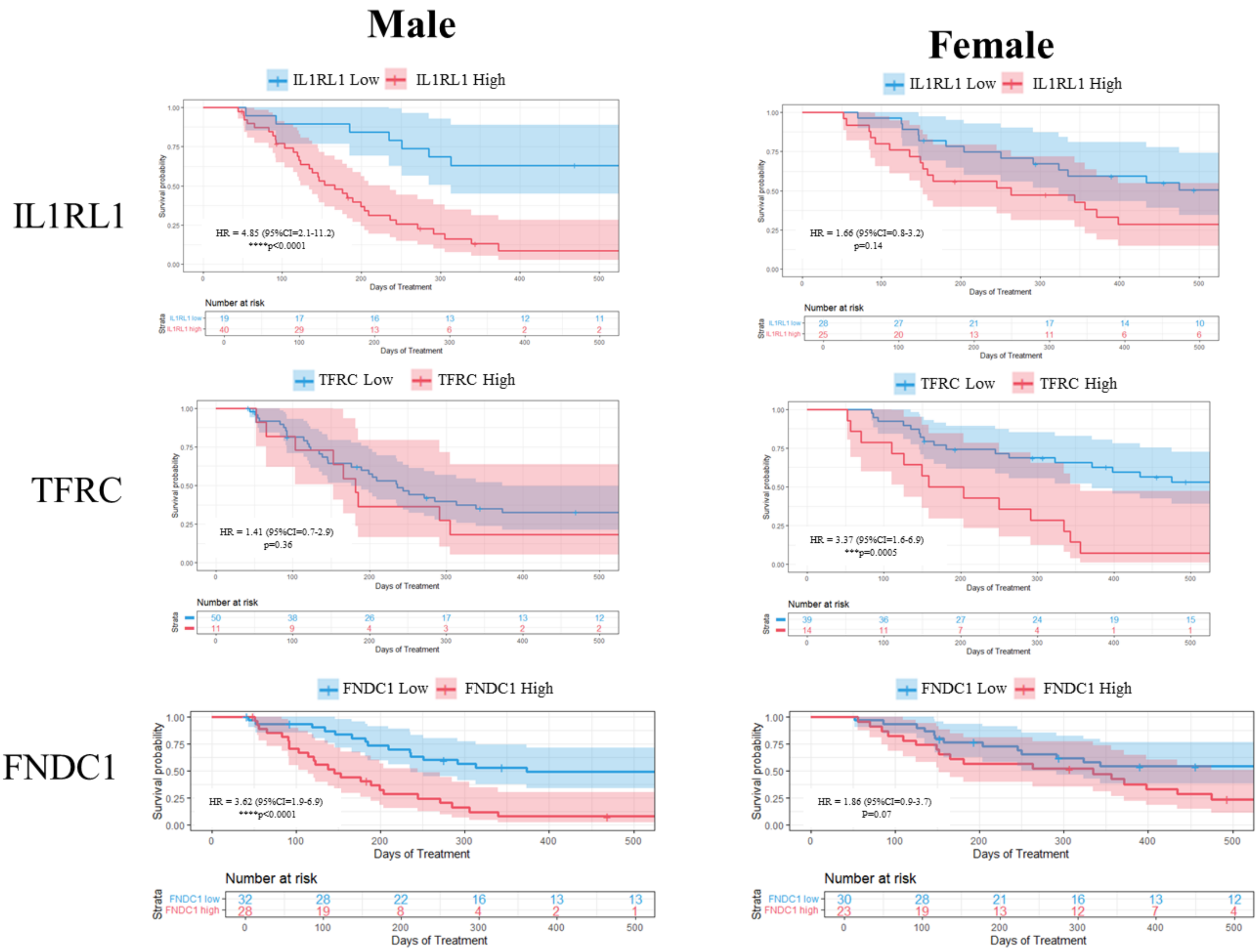
Male
Female
IL1RL1
TFRC
FNDC1
IL1RL1 Low
IL1RL1 High
HR = 4.85 (95%CI=2.1-11.2)
****p<0.0001
HR = 1.66 (95%CI=0.8-3.2)
p=0.14
TFRC Low
TFRC High
HR = 1.41 (95%CI=0.7-2.9)
p=0.36
HR = 3.37 (95%CI=1.6-6.9)
***p=0.0005
FNDC1 Low
FNDC1 High
HR = 3.62 (95%CI=1.9-6.9)
****p<0.0001
HR = 1.86 (95%CI=0.9-3.7)
P=0.07
Survival probability
Days of Treatment
Number at risk
Strata
FNDC1 low
FNDC1 high

**Supp. Fig. S5**

A)

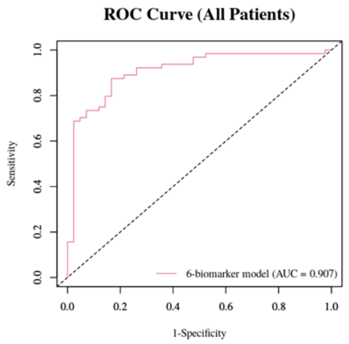


B)

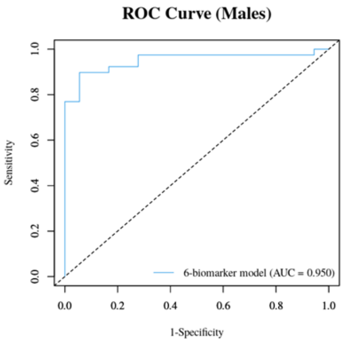


C)

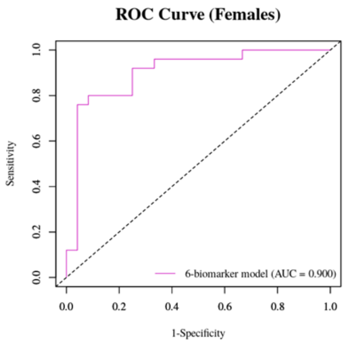


D)

| 6-Biomarker Model | **AUC (95% CI)** | **p-value** | **Sensitivity** | **Specificity** |
|---|---|---|---|---|
| All Patients | 0.907 (0.847-0.967) | $1.6x10^{-40}$ | 87.5% (76.8%-94.4%) | 83.3% (68.6%-93.0%) |
| Males | 0.950 (0.894-1.00) | $5.5x10^{-55}$ | 89.7% (75.8%-97.1%) | 94.4% (72.7%-99.9%) |
| Females | 0.900 (0.806-0.994) | $6.2x10^{-17}$ | 76.0% (54.9%-90.6%) | 95.8% (78.9%-99.9%) |
